# Efficacy of interventions to increase physical activity for people with heart failure: a meta-analysis

**DOI:** 10.1101/2021.04.14.21255358

**Authors:** Aliya Amirova, Theodora Fteropoulli, Paul Williams, Mark Haddad

## Abstract

**Objectives:** This meta-analysis aims to 1) evaluate the efficacy of physical activity interventions in heart failure and 2) to identify intervention characteristics significantly associated with the interventions’ efficacy.

**Methods:** Randomised controlled trials reporting intervention effects on physical activity in heart failure were combined in a meta-analysis using a random-effect model. Exploratory meta-analysis was performed by specifying the general approach (e.g. cardiac rehabilitation), strategies used (e.g. action planning), setting (e.g. centre-based), mode of delivery (e.g. face-to-face or online), facilitator (e.g. nurse), contact time, and behaviour change theory use as predictors in the random-effect model.

**Results:** Interventions (n=21) had a significant overall effect (*SMD* = 0.54,95% *CI*: [0.13; 0.95], *p* < 0.005). Combining an exercise programme with behaviour change intervention was found efficacious (*SMD* = 1.26, 95% *CI*: [0.26; 2.26], *p* < 0.05). Centre-(*SMD* = 0.98, 95% *CI*: [0.35; 1.62],*p* < 0.001), and group-based (*SMD* = 0.89, 95 % *CI*: [0.29; 1.50],*p* < 0.001) delivery by a physiotherapist (*SMD* = 0.84, 95% *CI*: [0.03; 1.65]], *p* < 0.01) were significantly associated with efficacy. The following strategies were identified efficacious: *prompts/cues* (*SMD* = 3.29, 95% *CI*: [1.97; 4.62]), credible source (*SMD*=2.08, 95% *CI*:[0.95;3.22]), *adding objects to the environment* (*SMD*=1.47, 95% *CI*:[0.41;2.53]), generalisation of the target behaviour (*SMD*=1.32, 95% *CI*:[0.22;2.41]), *monitoring of behaviour by others without feedback* (*SMD*=1.02, 95% *CI*:[0.05;1.98]), *self-monitoring of outcome(s) of behaviour* (*SMD*=0.79, 95% *CI*:[0.06;1.52]), *graded tasks* (*SMD*=0.73, 95% *CI*:[0.22;1.24]), *behavioural practice/rehearsal* (SMD=0.72, 95% *CI*:[0.26;1.18]), *action planning* (*SMD*=0.62, 95% *CI*:[0.03;1.21]), and *goal setting (behaviour)* (*SMD*=0.56, 95% *CI*:[0.03;1.08]).

**Conclusion:** The meta-analysis suggests intervention characteristics that may be suitable for promoting physical activity in heart failure. There is moderate evidence in support of an exercise programme combined with a behaviour change intervention delivered by a physiotherapist in a group- and centre-based settings.

**SUMMARY:** *What is already known about this subject?:* Individuals diagnosed with heart failure (HF) are advised to engage in physical activity. However, physical activity levels remain extremely low in this population group. Cardiac Rehabilitation (CR) is routinely offered to newly diagnosed HF patients. CR is multifaceted; It is unknown which specific components result in physical activity improvements once the programme has ended. It is essential to understand how best to improve everyday physical activity engagement in HF.

*What does this study add?:* This meta-analysis assessed what constitutes a successful physical activity intervention designed for individuals living with HF. The findings pinpoint specific intervention features and component that contribute to physical activity improvements in HF. Centre-based interventions that are delivered by a physiotherapist, in group format, that combine exercise with behaviour change intervention are promising for attaining physical activity improvements

*How might this impact on clinical practice?:* The findings of this meta-analysis may inform physical activity intervention designed for individuals diagnosed with HF. There is a need for additional training for physiotherapists in delivering behaviour change interventions alongside an exercise programme that includes the identified efficacious strategies. ***Note:*** all statistics are reported using Word Build-in: “Equation.”

## Introduction

The levels of engagement in physical activity of medically stable individuals diagnosed with heart failure (HF) are low [1]. Physical activity is a treatment strategy [2]. Cardiac rehabilitation (CR) and other exercise-based programmes have been shown to improve Quality of Life (QoL) [3,4] and reduce hospitalisation in HF [4,5]. However, a recent meta-analysis suggested that CR is less likely to be efficacious in sustaining physical activity in HF in particular compared to other cardiovascular diseases (CVD) [6]. The uptake of CR remains suboptimal [7]. Therefore, it is essential to evaluate the efficacy of alternative interventions as well as CR and identify content and features that are likely to be successful in promoting physical activity.

CR is a complex intervention. It is unclear which components are responsible for what outcomes and for which patient group [8]. There is a need to explore this intervention complexity and identify what makes an intervention successful [9]. Past reviews have suggested that short-term intervention effects are associated with strategies such as exercise prescription; goal setting; feedback and problem solving; and the use of a behaviour change theory [10].

CR might be missing some efficacious elements. Clark et al. (2014) pointed out that previous healthcare services research has not emphasised cardiac rehabilitation’s goal: how best to ensure that CVD patients benefit from a healthy lifestyle, including physical activity. Clark et al. (2014) also made a call to evaluate a range of potentially effective interventions that are facilitated by various professionals and make use of a diverse set of methods (e.g., remote monitoring). Evaluation of home-based and remote interventions is especially vital, given the recent restriction following the SARS-CoV-2 outbreak. It is also essential to understand what features of centre-based, group-based interventions contribute to physical activity improvement. The present meta-analysis of randomised controlled trials (RCTs) reviewed physical activity interventions, including CR, to identify intervention features that contribute to efficacy in improving physical activity.

## Methods

### Information sources

The review protocol was registered on PROSPERO database (CRD42015015280). Cochrane Library, MEDLINE, CINAHL, EMBASE, AMED, HEED, PsychARTICLES, PsychINFO, Global Health, Web of Science: Conference Proceedings, ‘Be Part of Research,’ and ClinicalTrials.gov were searched from inception to 20 February 2020. The search strategy is described in supplement 1.

### Eligibility criteria and study selection

Titles, abstracts, and full texts were independently screened by two reviewers (AA and PW). The criteria for considering RCTs were: i) adults diagnosed with HF, ii) intervention targeting physical activity (compared to usual care and/or education), and iii) report of a numerical result for physical activity outcome at intervention completion for both groups. Physical activity outcome was defined as any bodily movement produced by skeletal muscles that requires energy expenditure. Exercise is a subset of physical activity defined as structured physical activity [11]. Exercise, in the context of HF, is defined as selfcare behaviour (i.e., “I exercise regularly”).

### Data collection process

Relevant information was extracted from trial reports (article, supplementary materials, and protocols) using a standardised Cochrane data extraction form [12].

### Risk of bias in individual studies

The risk of bias at the study level was assessed using the Cochrane Collaboration Risk of Bias tool (2) [13] and informed sensitivity analysis.

### Data items

Interventions were classified in terms of their general approach to physical activity promotion (e.g. exercise), setting (e.g. home vs centre), mode of delivery (e.g. group vs individual), and facilitator (e.g. nurse). The Theory Coding Scheme (TCS) [14] was used to describe the extent to which trials employed a behaviour change theory in the intervention design. TCS scores range from 0 (no theory) to 8 (most extensive theory use). The intervention and comparator treatment were described in terms of the included behaviour change techniques. Interventions’ content was independently annotated by AA (100%) and TF (61.90%) using The Behaviour Change Techniques Taxonomy (BCTTv1) [15].

### Statistical analysis

Meta-analysis was performed using the `metafor` library in R [16]. A random effect model was used to estimate the overall efficacy of interventions using restricted maximum likelihood. The standardised mean difference (*SMD*) in physical activity levels between the main intervention and the comparator group was selected as the estimate of efficacy. Heterogeneity index (*I*^2^) was reported as the total unexplained variability in effect. Assessments at the 3-month, 6-month (short-term), and 12-month (long-term) follow-up were included. Meta-regression was performed to explore whether the efficacy was associated with the following: general approach (e.g. exercise programme), setting, mode of delivery (e.g. home-based), facilitator (e.g. nurse), behaviour change strategies (e.g. goal setting), and participant characteristics (i.e., mean age, NYHA class, proportion of males, mean EF (%), Aeschimic Aetiology (%)) were specified as predictors in the model [17]. We accounted for the fact that a small number of trials were presenting a particular intervention characteristic using Hartung-Knapp-Sidik adjustment as recommended by Debray et al. [19].

### Risk of bias across studies

The small study bias was evaluated using a funnel plot assessment and Egger’s test.

### Patient and public involvement

No patients were involved in formulating the research question, the outcome measures, or findings interpretation. Patients were not involved in planning or designing of the meta-analysis. This is due to the lack of funding available to include patients as partners in this meta-analysis. Results of this meta-analysis will be disseminated to the relevant patient organisations.

## Results

### Search results

Search results and reasons for exclusion are listed in the PRISMA diagram (Figure 1). A total of 20 trials evaluating 22 interventions post-completion (n = 21, [19–37]), at 6-months (n = 5, [29,32,33,35,38]), and 12-months (n = 5, [26,27,29,36–38]) follow-up were included in the meta-analysis.

**Figure 1.**
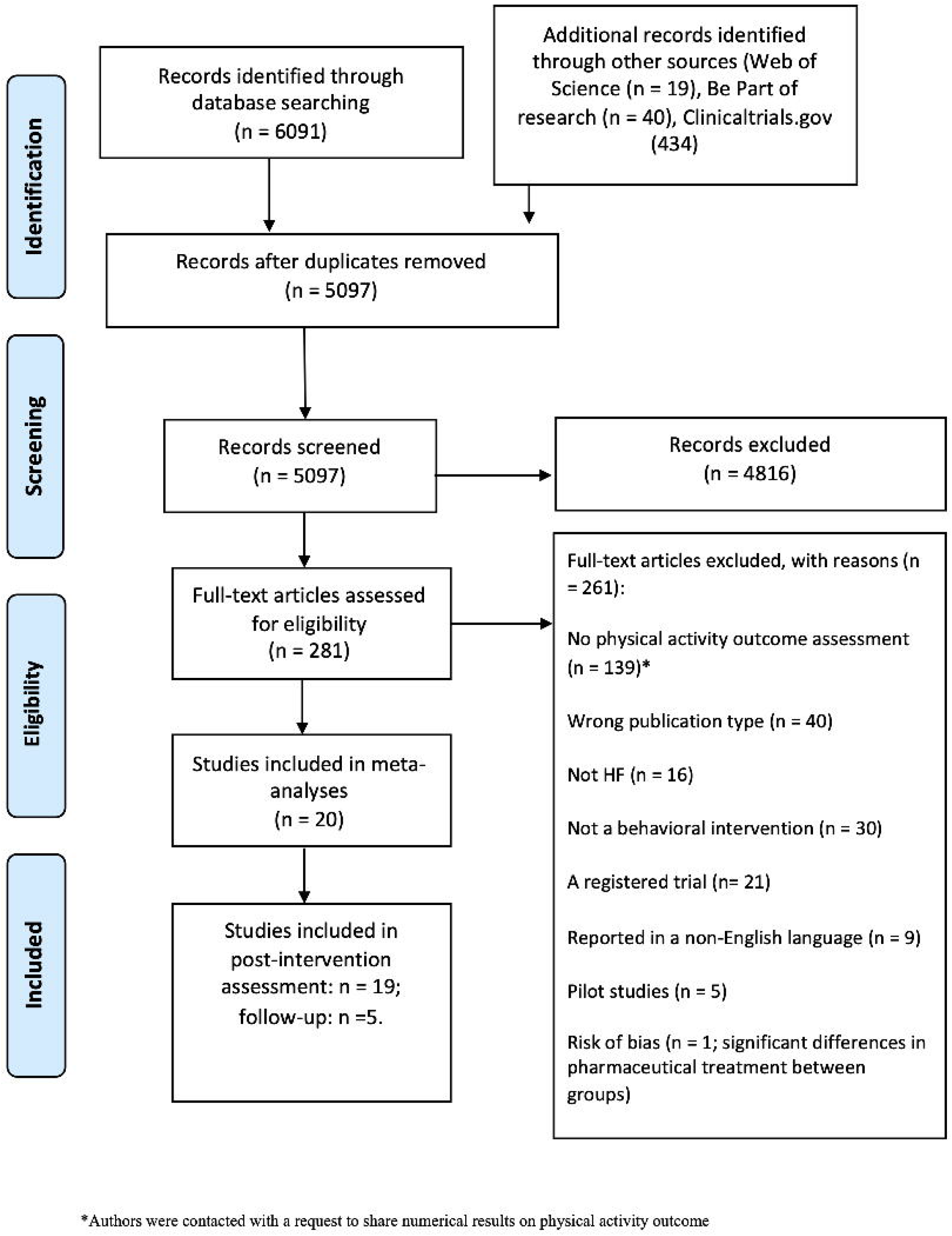
The study flow chart (PRISMA, 2009).

### Study characteristics

The trials were conducted between 1999 and 2018. The trials included a total of 6277 participants, and the median sample size was 100 [*IQR*: 60; 204]. A large proportion (37%) of participants were drawn from the HF-ACTION trial (n=2331)[37].

### Risk of Bias

The overall risk of bias is summarised in Figure 2. Six out of 20 trials reported low risk of bias [20,22,26,27,29,37]. A high risk of bias was present in two trials [19,30]. The sources of bias for each trial are summarised in supplement 2. Five trials evaluated the intervention against an active comparator: education [19,21,22,27,38]

**Figure 2.**
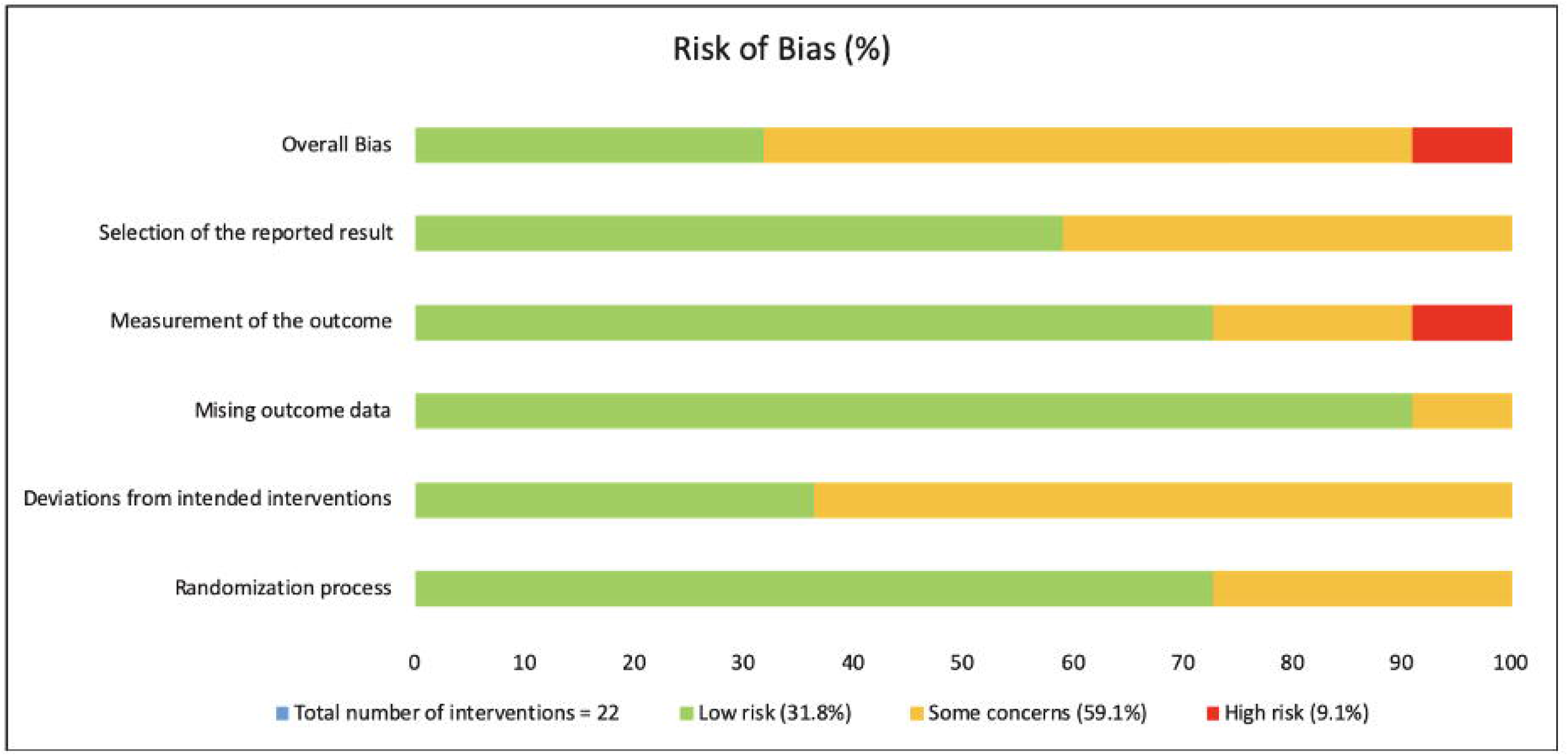
The risk of bias summary. The Cochrane Collaboration Risk of Bias tool (version 2) [15].

### Participant Characteristics

Mean age ranged from 54 [19] to 80 years old [33] (SD= 7.28; IQR:[62;70]), and the majority of the sample was male 69.49% (Table 1).

**Table 1.**
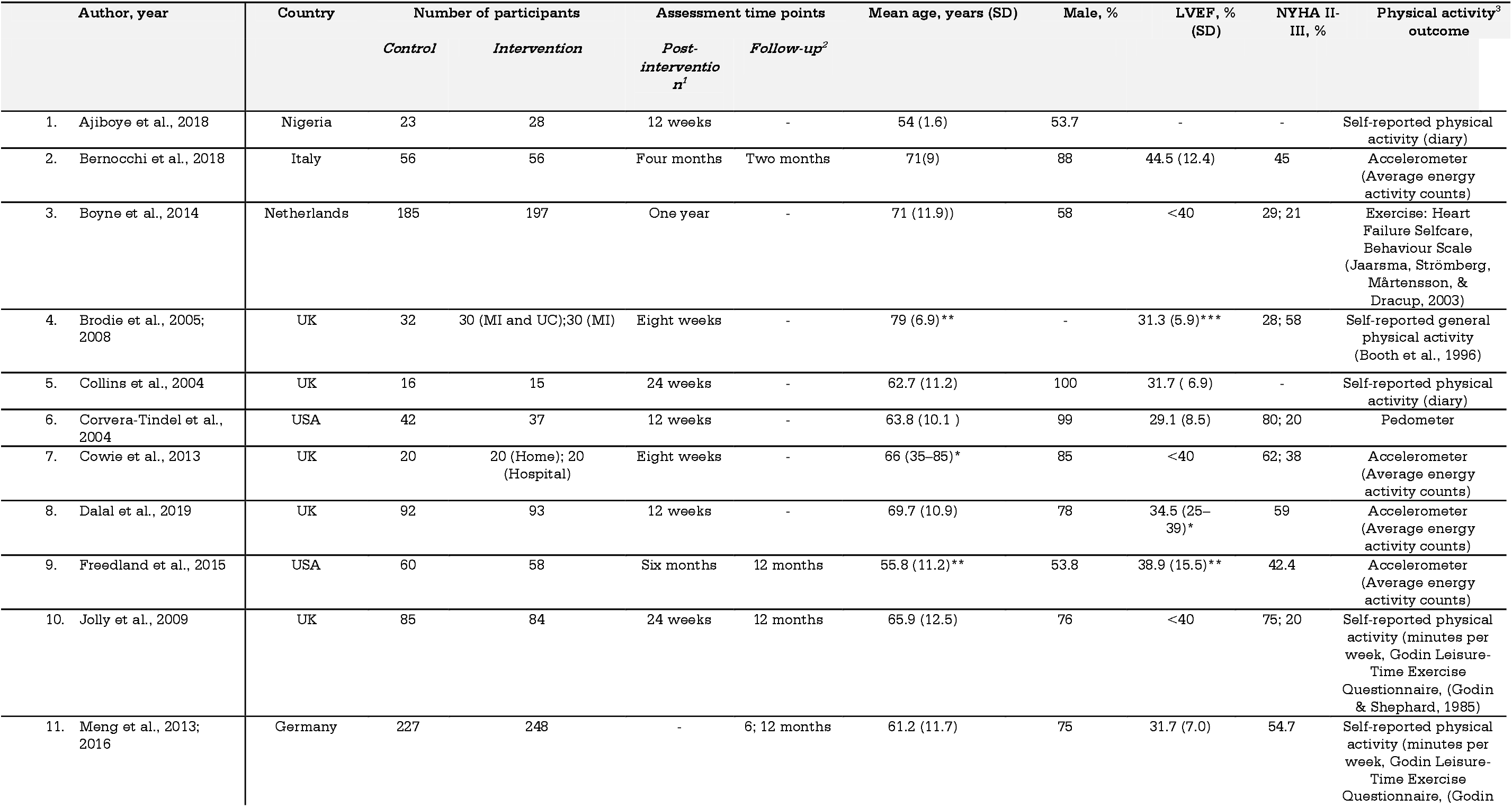
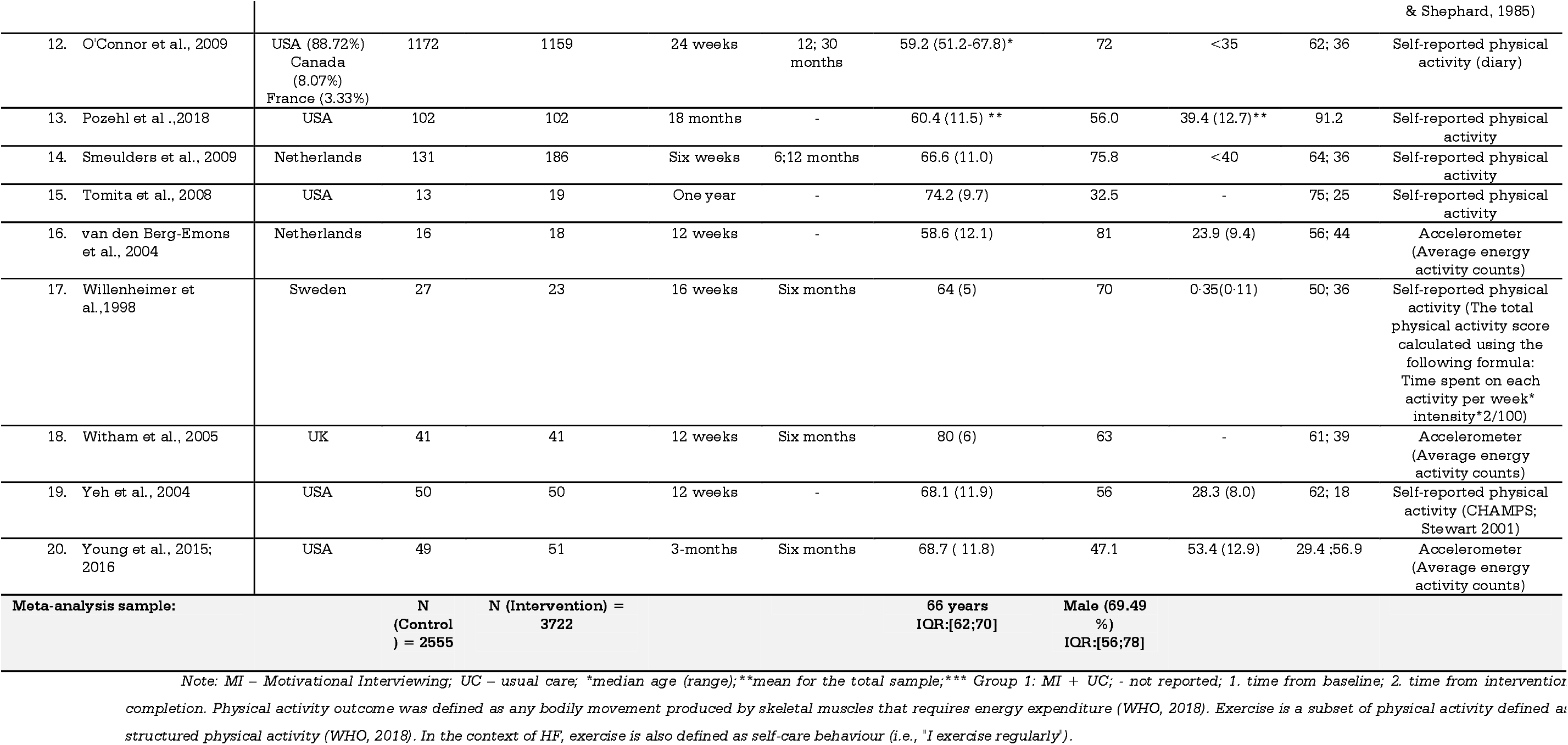
Study and Participant Characteristics.

### Post-completion efficacy

The present meta-analysis found a significant overall effect as assessed at post-completion (*SMD* = 0.54, 95% *CI*: [0.13; 0.95], *p* < 0.005). There was significant high heterogeneity in the estimated effect, *I*^2^ = 95.8%, (*Q* = 1531.74, *p* < 0.001) (Figure 3). The following intervention characteristics contributed to the heterogeneity in efficacy: general approach of the interventions, setting (i.e., centre-based vs home-based), facilitator, and several strategies.

**Figure 3.**
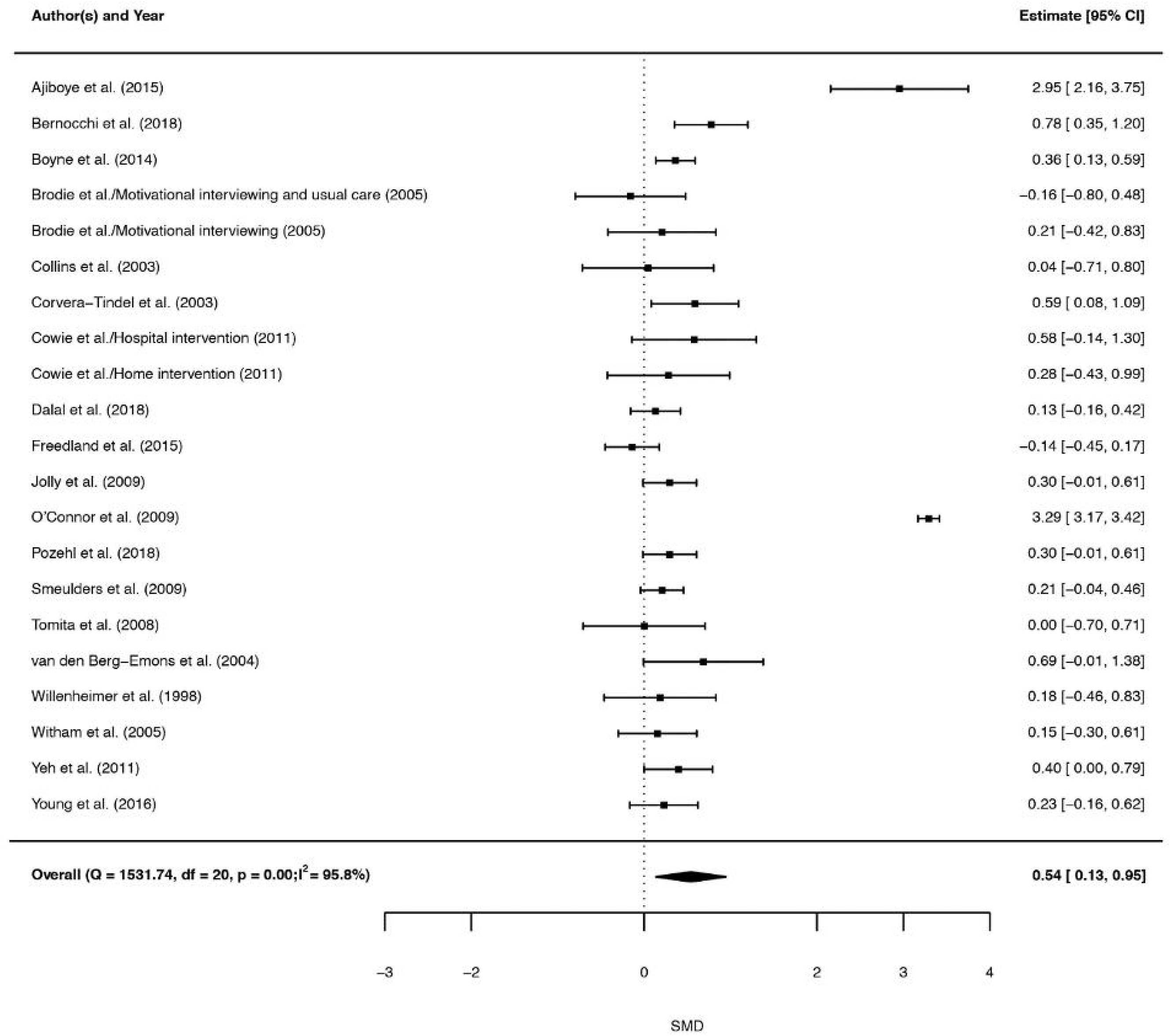
Forest plot illustrating overall estimated effect (*SMD*) and 95% *CI* and *SMD* and 95% CI for component trials.

### General approach

The included trials delivered interventions that were classified as *Exercise* (k=10, 47.62%), *Exercise and Behaviour Change* (k=3, 14.29%), *Motivational Interviewing* (k=2, 9.42%), *Remote Communication and Treatment* (k=3, 14.29%), *Cognitive Behavioural Therapy* (k=1, 4.76%), *Disease Management* (k=1, 4.76%), and *Self-Management* (k=1, 4.76%), Table 2. Exercise combined with behaviour change is an efficacious approach (Figure 4).

**Table 2.** Intervention Characteristics.

| Author, year |  | Intervention description provided by authors | General approach |
| --- | --- | --- | --- |
| Ajiboye et al., 2015 | *Main intervention* | Aerobic and resistance training and education | Exercise |
|  | *Comparator treatment* | usual care and education |  |
| Bernocchi et al., 2018 | *Main intervention* | Telerehabilitation and home-based personalised exercise maintenance programme | Remote Communication and Treatment |
|  | *Comparator treatment* | usual care |  |
| van den Berg-Emons et al., 2004 | *Main intervention* | Aerobic exercise training | Exercise |
|  | *Comparator treatment* | usual care without particular advice for exercise |  |
| Boyne et al., 2014 | *Main intervention* | Individually tailored e-health intervention 'Health Buddy' | Remote Communication and Treatment |
|  | *Comparator treatment* | education |  |
| Brodie et al., 2005 | *Main intervention 1* | Motivational Interviewing | Motivational Interviewing |
|  | *Main intervention 2* | Motivational Interviewing and education |  |
|  | *Comparator treatment* | education |  |
| Collins et al., 2004 | *Main intervention* | Aerobic exercise training | Exercise |
|  | *Comparator treatment* | usual care |  |
| Corvera-Tindel et al., 2004 | *Main intervention* | A home walking exercise programme | Exercise |
|  | *Comparator treatment* | usual care |  |
| Cowie et al., 2011 | *Main intervention 1* | Hospital-based aerobic exercise training | Exercise |
|  | *Main intervention 2* | home-based exercise training |  |
|  | *Comparator treatment* | usual care |  |
| Dalal et al., 2019 (REACH-HF) | *Main intervention* | Rehabilitation enablement in HF: self-care and rehabilitation | Exercise and Behaviour Change |
|  | *Comparator treatment* | usual care |  |
| Freedland et al., 2018 | *Main intervention* | Integrative Cognitive Behaviour Therapy and education and usual care | Cognitive Behaviour Therapy |
|  | *Comparator treatment* | Enhanced (with education) usual care |  |
| O'Connor et al., 2009 (HF-ACTION) | *Main intervention* | Aerobic exercise training and Exercise adherence facilitation intervention | Exercise and Behaviour Change |
|  | *Comparator treatment* | usual care |  |
| Jolly et al., 2009 | *Main intervention* | Aerobic and resistance exercise training | Exercise |
|  | *Comparator treatment* | HF specialist nurse care |  |
| Meng et al., 2016 | *Main intervention* | Self-management patient education program and CR | Self-Management |
|  | *Comparator treatment* | education |  |
| Pozehl et al., 2018 (HEART Camp) | *Main intervention* | Multicomponent intervention and resistance exercise training | Exercise and Behaviour Change |
|  | *Comparator treatment* | Enhanced (nine exercise sessions for three months) |  |
| Smeulders et al., 2009 | *Main intervention* | Chronic disease management programme | Disease |
|  |  |  | Management |
|  | *Comparator treatment* | usual care |  |
| Tomita et al., 2008 | *Main intervention* | Multidisciplinary internet-based programme on HF management | Remote Communication and Treatment |
|  | *Comparator treatment* | usual care |  |
| Willenheimer et al., 2001 | *Main intervention* | Aerobic exercise training | Exercise |
|  | *Comparator treatment* | usual care and discouragement to exercise |  |
| Witham et al., 2005 | *Main intervention* | Seated aerobic and resistance exercise training | Exercise |
|  | *Comparator treatment* | usual care |  |
| Yeh et al., 2011 | *Main intervention* | Exercise training (Tai Chi Mind-Body movement) | Exercise |
|  | *Comparator treatment* | usual care |  |
| Young et al. 2015; 2016 | *Main intervention* | Patient Activation Intervention on self-management in HF | Self-Management |
|  | *Comparator treatment* | usual care |  |

**Figure 4.**
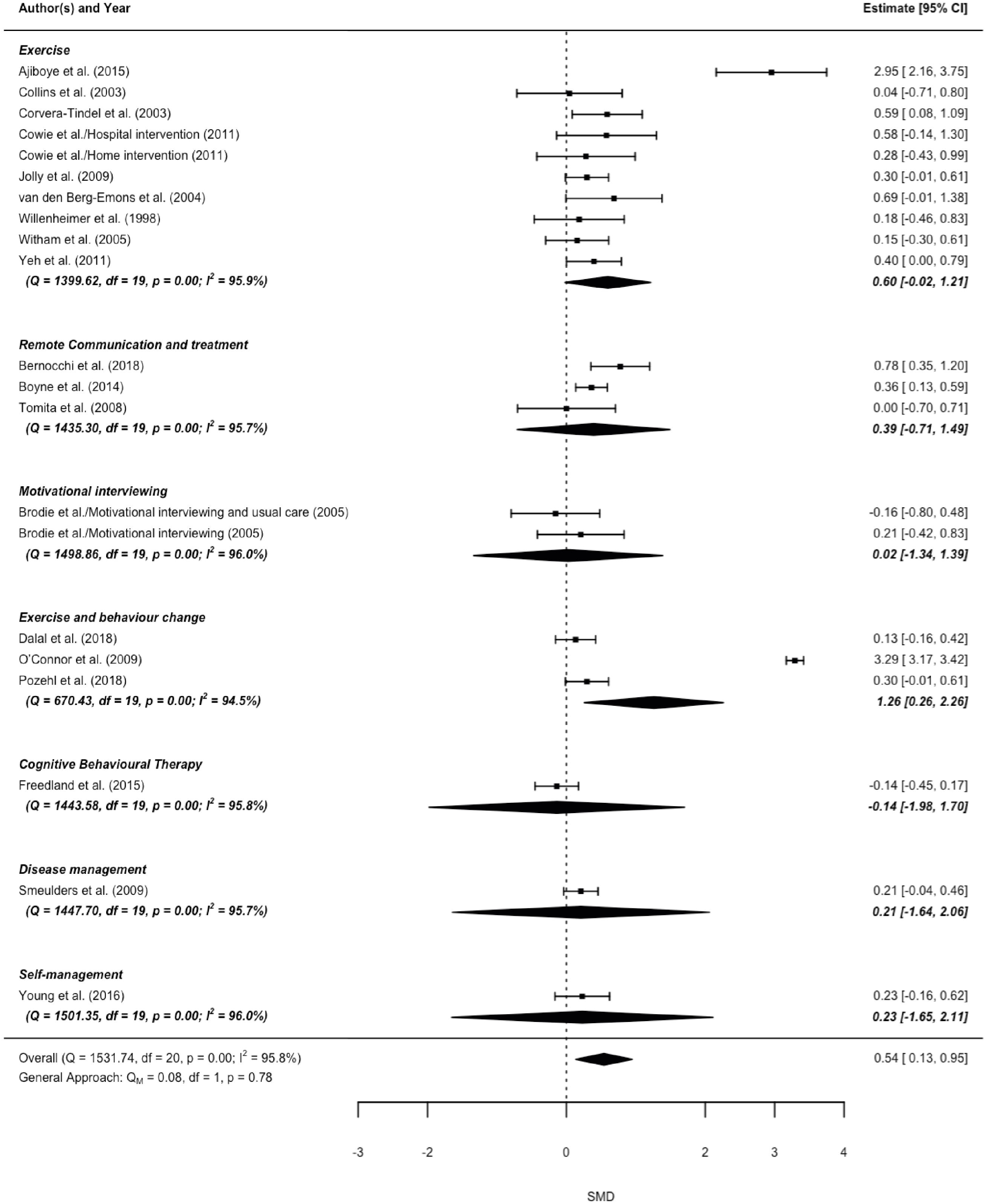
Forest plot illustrating the standardised mean differences (*SMD*, 95 % *CI*) moderated by the general approach.

### Intervention strategies

A total of 38 strategies (i.e., BCTs) were present across included trials (supplement 4). Interventions included a mean of 8.90 (*SD* = 3.77; *IQR* = [8;10]) strategies. The following strategies were associated with moderate to large effects: *prompts/cues, credible source, adding objects to the environment, generalisation of target behaviour, monitoring of behaviour by others without feedback, self-monitoring of outcome(s) of behaviour, graded tasks, behavioural practice/rehearsal, action planning, goal setting* (*behaviour*) (*SMD*: 0.56-3.29, Table 3).

**Table 3.** Intervention Characteristics Associated with Efficacy.

| Intervention Characteristics | SMD | CI, 95% |
| --- | --- | --- |
| ***Behaviour Change Techniques:*** |  |  |
| Prompts/cues<br>Definition: Introduce or define environmental or social stimulus to promote or cue the behaviour. Examples: frequent phone calls by a health professional/ post or email reminders | 3.29 | [1.97;4.62] |
| Credible source<br>Definition: present verbal or visual communication from a credible source in favour of or against the behaviour. Examples: Explicit, detailed and salient advice from a health professional to engage in physical activity. | 2.08 | [0.95;3.22] |
| Adding objects to the environment<br>Definition: Add objects to the environment in order to facilitate the performance of the behaviour. Examples: Provision of a treadmill, weights, step, or stationary bicycle. | 1.47 | [0.41;2.53] |
| Generalisation of the target behaviour<br>Definition: Advice to perform the desired behaviour, which is already performed in a particular situation, in another situation. Examples: Encouragement to engage in an exercise in home settings. | 1.32 | [0.22;2.41] |
| Monitoring of behaviour by others without feedback<br>Definition: Observe or record behaviour with the person's knowledge as part of a behaviour change strategy. Examples: The physiotherapist informs participants that their physical activity levels will be monitored using accelerometers and telemonitoring devices. | 1.02 | [0.05;1.98] |
| Self-monitoring of outcome(s) of behaviour<br>Definition: Establish a method for the person to monitor and record the outcome(s) of their behaviour as part of a behaviour change strategy. Examples: Monitoring reduced pain symptoms and dyspnoea as a result of physical activity. | 0.79 | [0.06;1.52] |
| Graded tasks<br>Definition: Set easy-to-perform tasks, making them increasingly difficult, but achievable until the behaviour is performed. Examples: Gradual increase in the level of exertion as assessed using the Borg scale. | 0.73 | [0.22;1.24] |
| Behavioural practice/rehearsal<br>Definition: Prompt practice or rehearsal of the performance of the behaviour one or more times in a context or at a time when the performance may not be necessary. Examples: Exercise training (individual or in a group). | 0.72 | [0.26;1.18] |
| Action planning<br>Definition: prompt, detailed planning of performance of the behaviour (must include at least one of context, frequency, duration and intensity). Examples: plan when, where, how much and at what intensity the participant will perform the exercise. | 0.62 | [0.03;1.21] |
| Goal setting (behaviour)<br>Definition: set or agree on a goal defined in terms of the behaviour to be achieved. Examples: Set a goal to complete 30 minutes of exercise (brisk walking) at the vigorous intensity in future. | 0.56 | [0.03;1.08] |
| ***Setting:*** Centre-based interventions | 0.98 | [0.35;1.62] |
| ***Mode of delivery:*** Group-based interventions | 0.89 | [0.29;1.50] |
| ***Facilitator:*** Physiotherapist | 0.84 | [0.03;1.65] |

Note: Definitions are from Michie et al. [15]. Intervention characteristics are described in Table 2 and supplement 4. SMD and 95% CI for characteristics that were not suggested to be significantly associated with efficacy are summarised in supplement 4.

### Settings, facilitator, and duration

Interventions were delivered at home (n = 8, 38%), in a hospital/clinic (n = 8, 38%), or both (n = 5, 24%). Only centre-based delivery significantly moderated the efficacy of the included interventions, Table 3. Interventions were facilitated by general practice nurses (n = 9, 42.85%), physiotherapists (n = 6, 28.6%), HF nurses (n = 4, 19%), exercise instructors (n = 3, 14.29%), researchers (n = 2, 9.42%), lay leaders (n = 1, 4.76%), advanced practice nurse (n = 1, 4.76%), psychologists (n = 14.76%), and clinical psychology trainees (n = 1, 4.76%). Intervention delivery by a physiotherapist was associated with efficacy (Table 3). Intervention duration varied from one day to 72 weeks. Mean contact time was 1849.38 minutes (SD = 1716.40) and was not associated with intervention efficacy.

### Theory use

Seven interventions were based on a behaviour change theory (supplement 4). The *extent* of theory use (TCS) was not associated with efficacy (SMD=0.13, p = 0.059, CI: [-0.006; 0.27]).

Sample characteristics, including mean age, gender, mean LVEF (%), NYHA class, and aetiology, were not significantly associated with intervention efficacy (supplement 5). Likewise, the differences in efficacy between trials using self-reports and trials using accelerometer or pedometer were non-significant (supplement 5).

### Long-term efficacy

The included interventions assessed physical activity at a 2-month, 6-month, 12-month and 30-months follow-up. The overall short-term effect was non-significant at the 6-month, *SMD* = −0.06 (95% *CI*: [−0.49; 0.38], *p* = 0.80) and 12-month follow-up, *SMD* = −0.11 (*p* = 0.74, 95% *CI*: [−0.77; 0.55]). Due to the small number of interventions reporting follow-up assessment, it was not feasible to evaluate the long-term effects associated with the individual intervention characteristics.

### Sensitivity Analysis

Interventions were compared to usual care [20,23–26,28–30,33–37], education delivered by a HF specialist nurse [21,22,27,38] or unspecified health professional [19], and discouragement to exercise [32]. The comparator treatments included a mean of 1.15 (*SD* = 1.49) strategies (supplement 4). When trials comparing the main intervention to education were excluded, the effects of *Exercise and Behaviour Change, Remote Monitoring and Treatment*, and *Exercise* were significant (supplement 6). The exclusion of a large (N= 2331) trial with a younger (56 years old) sample [37] resulted in a significant decrease in the overall effect. The effect estimates for *Exercise* and *Behaviour Change* approach, *prompts/cues, credible source, adding objects to the environment, generalisation of the target behaviour, monitoring of behaviour by others without feedback, self-monitoring of outcome(s) of behaviour, action planning, and goal setting (behaviour)* were sensitive to the inclusion of the trial (supplement 6). The exclusion of interventions with a high risk of bias indicated that the efficacy of *Exercise* approach was overestimated. The effects of the following strategies were underestimated: *social support (emotional), social support (practical), theory use (TCS score), information about health consequences, and information on how to perform the behaviour*.

### Small study bias

A funnel plot for SMD against standard error is available in supplement 7. The Egger’s test suggested a lack of publication and small study bias (test for funnel plot asymmetry: *Z* = −0.46, *p* = 0.64).

## Discussion

The present meta-analysis found moderate evidence in support of existing physical activity interventions designed for individuals living with HF. Centre-based interventions that are delivered by a physiotherapist, in group format, that combine exercise with behaviour change intervention are promising for attaining physical activity improvements. Intervention strategies identified as efficacious are: *prompts/cues, credible source, adding objects to the environment, generalisation of the target behaviour, monitoring of behaviour by others without feedback, self-monitoring of outcome(s) of behaviour, graded tasks, behavioural practice/rehearsal, action planning, and goal setting (behaviour)*. To our knowledge, this is the first meta-analysis evaluating the components of behavioural interventions that are associated with increased physical activity in HF. Interventions that were delivered by a physiotherapist in a centre-based setting were more promising in attaining physical activity improvement than home-based interventions or those delivered by facilitators other than physiotherapist (i.e., nurse, lay leader, researcher). This is in contrast to the findings of a previous meta-analysis suggesting that centre-based and home-based programmes delivered to individuals post-myocardial infarction or revascularisation, and with HF are equivalent in their efficacy in improving survival, QoL, and exercise capacity [39]. The present meta-analysis found that delivery of an intervention to a group contributed to efficacy. However, given the ongoing pandemic, it is essential to optimise delivery of physical activity interventions in home settings. Group-based interventions contribute to behavioural change via social comparison, changes in normative beliefs about health behaviour, and group member identity [40]. These factors can also be considered when designing home-based, contact-free physical activity interventions for older adults with HF.

A previous systematic review of CR programmes found that, in general, educational and behavioural elements of CR did not result in physical activity improvements beyond those achieved by exercise-based programmes [6]. However, behavioural elements are diverse and vary in their efficacy. The present meta-analysis evaluated a range of such elements and outlined those that are efficacious. A combination of an exercise and behaviour change approach was found to be more efficacious than other approaches, including exercise alone. Several strategies to improve physical activity appear promising (Table 3). Theoretical explanations for the efficacy of these strategies were previously offered [41]. *Graded tasks* exert an effect on physical activity by fostering positive beliefs about capability through skill mastery (e.g. exercise training) [41]. *Self-monitoring, monitoring by others, planning, goal-setting, and feedback* are theorised to improve control and regulation of behaviour [42]. Finally, the efficacy of adding an object associated with physical activity (e.g. treadmill) indicates the relevance of cueing (i.e., automatic association and non-deliberate regulation of behaviour) [41].

### Implications for clinical practice and future research

The present meta-analysis found moderate evidence in support of combining exercise programme with behaviour change intervention, delivered by a physiotherapist. Thus, there is a need for additional training for physiotherapists in delivering behaviour change interventions that will include the identified efficacious strategies. Practical limitations of the identified efficacious strategy need to be considered when designing interventions. *Adding objects to the environment* to support physically activity lifestyle (e.g. a treadmill) may not be affordable or practical, and does not satisfy the principle of health equity [43]. In addition, further research investigating how best to promote a physically active lifestyle in the older HF population is encouraged. The clinical profiles of older adults differ from younger adults, with a significantly worse prognosis and a larger number of comorbidities in the former [44]. Older adults may also differ in their beliefs about physical activity; and strategies that are suited for promoting an active lifestyle in older adults are different to those that are efficacious for the general population [45]. Investigation of which behaviour change theory should form the basis for an intervention is also warranted. Only five trials assessed physical activity at 6- and 12-month follow-ups. Long-term efficacy was not supported. Thus, it is important to investigate how sustained physical activity improvements can be established.

### Study-level limitations

High risk of bias was observed in two trials [21,32]. The sensitivity analysis indicated that the inclusion of these trials may overestimate the efficacy of exercise programmes and underestimate the efficacy of remote monitoring and treatment. Remote communication and feedback interventions that include strategies such as biological feedback (e.g. symptom monitoring and feedback) delivered by a nurse using telehealth device, as well as self-monitoring of the behaviour, and information about health consequences [22,23] are identified as efficacious when high risk of bias trials are excluded. The HF-ACTION [37] trial constituted the majority of the meta-analysis sample and when it was excluded in the sensitivity analysis, only a small non-significant effect of exercise combined with behaviour change was observed. High-quality trials assessing the short and long-term effects of behaviour change; remote communication and treatment; and exercise programmes on physical activity in older adults (>70 years old) with HF are required.

### Strengths and limitations of the review

A Cochrane overview of reviews recommended exploring intervention complexity using meta-regression to evaluate the association between intervention characteristics and efficacy [9]. This meta-analysis identified, annotated, and classified behaviour change interventions in terms of their general approach, strategies, settings, facilitator, delivery mode, duration, and use of theory; and using meta-regression assessed the association between these characteristics and the efficacy. The clear, consistent, and systematic description of the interventions facilitated the reliable grouping and analysis. This helped pinpoint specific efficacious features and elements that can be applied, either as part of CR or otherwise, to improve physical activity outcomes in HF. However, there are a few limitations. Intervention features were present in clusters across the included trials. Given the small number of RCTs evaluating any single included characteristic, multiple comparisons were not feasible. It is not possible to ascertain whether each of the evaluated features is efficacious on their own or only in combination. These features need to be evaluated in a multi-arm trial comparing their effects.

## CONCLUSIONS

This meta-analysis explored intervention complexity and identified some features of potentially promising physical activity interventions designed for people living with HF. The present review provides moderate evidence that an exercise programme combined with a behaviour change intervention is a promising approach to increasing physical activity in HF. The meta-analysis suggests behaviour change strategies that may be useful in promoting physical activity in HF.

## Data Availability

Data is available upon request

## Acknowledgements

We would like to thank Professor Gary Morgan at City, University of London for his support throughout the review. We also would like to thank the authors of the included studies for providing information required for the detailed description of the interventions and data-analysis. We also thank Dr Michael J. Shoemaker (Grand Valley State University) for sharing details on one of the pilot studies he and his colleagues carried out (excluded at the later stage of the review).

## Contributors

All authors contributed to design and planning. AA and PW contributed by screening articles for inclusion. AA and TF contributed to data extraction and risk of bias assessment. AA implemented the analysis. AA, TF, and MH contributed to reporting. TF, MH contributed to the critical revision of the manuscript.

## Patient consent

Not required.

## Ethics committee approval

Not required.

## Supplementary files

Supplement 1: Search strategy.

Supplement 2: Figure. Risk of bias (individual studies).

Supplement 3: Figure. The dispersion (tau) of the underlying main effect.

Supplement 4. Expanded Table 2: Intervention characteristics.

Supplement 5. Nonsignificant results (exploratory meta-analysis).

Supplement 6: The sensitivity analysis results.

Supplement 7: Funnel plot.

