## Supplement 1 for "Efficacy of interventions to increase physical activity for people with heart failure: a meta-analysis"

**Supplement 1: Search strategy**

Interface - EBSCOhost Research Databases
Search Screen - Advanced Search

Database - Academic Search Complete; Art Full Text (H.W. Wilson); Business Source Complete; CINAHL Complete; Communication Source; Criminal Justice Abstracts with Full Text; eBook Collection (EBSCOhost); eBook Nursing Collection; EconLit with Full Text; E-Journals; European Views of the Americas: 1493 to 1750; GreenFILE; Health Policy Reference Center; Library, Information Science & Technology Abstracts; Library, Information Science & Technology Abstracts with Full Text; MathSciNet via EBSCOhost; MEDLINE Complete; Political Science Complete; APA PsycArticles; APA PsycBooks; APA PsycInfo; APA PsycTests; Regional Business News;RILM Abstracts of Music Literature (1967 to present); SocINDEX with Full Text; Teacher Reference Center.

Cochrane Library, MEDLINE, CINAHL, EMBASE, AMED, HEED, PsychARTICLES, PsychINFO, Global Health, Web of Science: Conference Proceedings, ‘Be Part of Research,’ and ClinicalTrials.gov.

**Search Strategy (via EBSCOhost)**

(MM "Heart Failure+")

"heart failure"

(MM "Cardiac Output, Decreased")

(MM "Ventricular Dysfunction+")

heart N5 fail*

cardi* N4 dysfunction*

heart N5 dysfunction*

"congestive heart failure"

"cardiac fail*"

"systolic heart failure"

"cardiac incompetence"

"cardiac decompensation"

"cardiac insufficiency"

"chronic heart failure"

"cardial insufficiency"

"myocardial failure"

"myocardial insufficiency"

"heart N3 fail*"

"diastolic dysfunction*"

"Systolic dysfunction*"

TITLE: Efficacy of interventions to increase physical activity for people with heart failure: a meta-analysis

"heart N3 dysfunction*"

"cardiac dysfunction*"

OR/1-22

(MH "Behavioral Changes")

(MH "Life Style Changes")

(MM "Self Care+")

"Self-management"

“Intervention”

(MM "Early Intervention+")

(MM "Patient Care+")
(MM "Rehabilitation+")
(MM "Home Rehabilitation+")
(MM "Rehabilitation, Cardiac+")
(MM "Rehabilitation, Community-Based")
(MH "Physical Education, Adapted")
(MH "Behavioral Objectives")
(MH "Psychosocial Adjustment: Life Change (Iowa NOC)
(MH "Change Management")
(MH "Behavior Management (Iowa NIC)")
(MH "Health Behavior")
(MH "Phychotherapy+")
"Behavio$ral intervention"
"Behavio$change technique*"
"Behavio$r change"
"Counselling"
"Psychotherapy"
OR/24-46
(MH "Physical Activity")
(MH "Sports+")
(MH "Activities of Daily Living+")
(MH "Exercise+")
(MH "Leisure Activities+")
(MH "Physical Fitness+")
(MH "Movement")
(MH "Aerobic Exercise+")
(MH "Swimming")
(MH "Rehabilitation, Cardiac")
(MH "Resistance training")
(MH "Sports Specific Training")
(MH "Group Exercise")
Physical N5 activ*
Exercis*
OR/48-62
(MH "Randomized Controlled Trials")
"Randomi$ed controlled trial"
"Clinical trial"
OR/64-66
23 AND 47
23 AND 47 AND 63
23 AND 47 AND 63 AND 67

TITLE: Efficacy of interventions to increase physical activity for people with heart failure: a meta-analysis

### Search Strategy (via OVID):

1. exp heart failure/
2. heart failure.mp.
3. heart decompensation.mp.
4. heart insufficiency.mp.
5. cardiac failure.mp.
6. cardiac incompetence.mp.
7. cardiac decompensation.mp.
8. cardiac insufficiency.mp.
9. exp heart output/
10. cardiac output.mp.
11. exp diastolic dysfunction/
12. exp congestive heart failure/
13. diastolic dysfunction.mp.
14. exp systolic dysfunction/
15. exp heart left ventricle failure/
16. heart left ventricle failure.mp.
17. cardial insufficiency.mp.
18. chronic heart failure.mp.
19. chronic heart insufficiency.mp.
20. decompensation,heart.mp.
21. myocardial failure.mp.
22. myocardial insufficiency.mp.
23. (heart adj3 fail*).tw.
24. (heart adj3 dysfunction*).tw.
25. left ventricular dysfunction.tw.
26. (cardiac adj3 dysfunction*).tw.
27. (cardiac adj3 fail).tw.
28. ventricular dysfunction.mp.
29. chronic cardiac failure.mp.
30. congestive cardiac failure.mp.
31. 1 or 2 or 3 or 4 or 5 or 6 or 7 or 8 or 9 or 10 or 11 or 12 or 13 or 14 or 15 or 16 or 17 or 18 or 19 or 20 or 21 or 22 or 23 or 24 or 25 or 26 or 27 or 28 or 29 or 30
32. exp physical activity/
33. physical exercise.mp.
34. physical activity.mp.
35. exercise.mp.
36. exp aerobic exercise/
37. aerobic exercise.mp.
38. exp resistance training/
39. resistance training.mp.
40. exercise training.mp.
41. exp daily life activity/
42. exp walking/
43. exp motor activity/
44. daily physical activity.mp.
45. exp motor activity/
46. exp leisure/
47. leisure activities.mp.
48. exp heart rehabilitation/
49. cardiac rehabilitation.mp.
50. exercise program.mp.
51. exercise programme.mp.
52. exp fitness/
53. exp swimming/
54. exp sport/
55. exp endurance training/
56. (physic* adj3 activ*).tw.

57. physical activity.tw.
58. exercis*.tw.
59. walk*.tw.
60. (daily adj5 physic adj5 activ*).tw.
61. 32 or 33 or 34 or 35 or 36 or 37 or 38 or 39 or 40 or 41 or 42 or 43 or 44 or 45 or 46 or 47 or 48 or 49 or 50 or 51 or 52 or 53 or 54 or 55 or 56 or 57 or 58 or 59 or 60
62. 31 and 61
63. exp intervention study/
64. intervention.mp.
65. exp health promotion/
66. exp behavior change/
67. behavioral intervention.mp.
68. behaviour change.mp.
69. exp behavior change/
70. psychological intervention.mp.
71. exp patient education/
72. exp counseling/
73. exp patient counseling/
74. behav* change.tw.
75. (change adj3 behavio$r).tw.
76. intervention.tw.
77. health promotion.tw.
78. behavio$r change technique*.tw.
79. behavio$r change strateg*.tw.
80. BCT.tw.
81. randomized controlled trial.tw.
82. randomized controlled trial/
83. clinical trial/
84. controlled study/
85. RCT.mp.
86. 81 or 82 or 83 or 84 or 85
87. cardiac rehabilitation.tw.
88. 63 or 64 or 65 or 66 or 67 or 68 or 69 or 70 or 71 or 72 or 73 or 74 or 75 or 76 or 77 or 78 or 79 or 80 or 87
89. 31 and 61 and 86 and 88
