## Supplement 2 for "Efficacy of interventions to increase physical activity for people with heart failure: a meta-analysis"

| Study ID | Experimental | Comparator | Randomization process | Deviations from intended interventio | Missing outcome data | Measurement of the outcome | Selection of the reported result | Overall |
|---|---|---|---|---|---|---|---|---|
| Ajiboye et al. 2015 | Aerobic and reisistnace and education traning | Education | + | ? | + | - | ? | - |
| Bernocchi et al. 2018 | Telerehabilitation with personalised exercise | Usual care | + | ? | + | + | + | + |
| Boyne et al. 2015 | Individually tailored e-health intervention | Education | + | ? | + | + | ? | ! |
| Brodie et al. 2005 | Motivational interviewing and education | Education | + | ? | + | + | + | + |
| Brodie et al. 2005 | Motivational interviewing | Education | + | ? | + | + | + | + |
| Collins et al. 2004 | Aerobic exercise training | Usual care | ? | ? | + | + | ? | ! |
| Corvera-Tindel et al. 2004 | Aerobic exercise training | Usual care | ? | + | + | + | + | ! |
| Cowie et al. 2013 | Home-based exercise training | Usual care | ? | ? | + | + | + | ! |
| Cowie et al. 2013 | Hospital-based exercise training | Usual care | ? | ? | + | + | + | ! |
| Dalal et al. 2018 | The rehabilitaiton enablement in chronic heart failure | Usual care | + | + | + | + | + | + |
| Freedland et al. 2015 | Cognitive Behavioural Therapy | Education | + | + | + | + | + | + |
| Jolly et al. 2009 | Aerobic and resistance exercise care | Usual care | + | ? | + | ? | ? | ! |
| Meng et al. 2016 | Self-management patient education program | Education | ? | + | ? | ? | + | ! |
| O'Connor et al. 2009 | Aerobic exercise training exercise adherance intervention | Usual care | + | + | + | + | + | + |
| Pozehl et al. 2018 | Multicomponent intervention with exercise | Usual care | ? | ? | + | ? | + | ! |
| Smeulders et al. 2009 | Chronic disease manamgent | Usual care | + | + | + | + | + | + |
| Tomita et al. 2008 | Multidisciplinary internet program | Usual care | + | ? | ? | - | ? | - |
| Van den Berg-Emons et al. 2004 | Aerobic exercise training | Usual care | + | ? | + | + | ? | ! |
| Wellenheimer et al. 1998 | Aerobic exercise training | Discouragement to exercise | + | ? | + | ? | ? | ! |
| Witham et al. 2005 | Seated aerobic exercise | Usual care | + | + | + | + | ? | ! |
| Yeh et al. 2004 | Tai-Chi mind body exercise | Usual care | + | + | + | + | ? | ! |
| Young et al. 2015 | Patient activation programme on self-management | Usual care | + | ? | + | + | + | ! |

+ Low risk
? Some concerns
- High risk
