## Supplementary figures and images for "Efficacy of interventions to increase physical activity for people with heart failure: a meta-analysis"

### Supplement 3

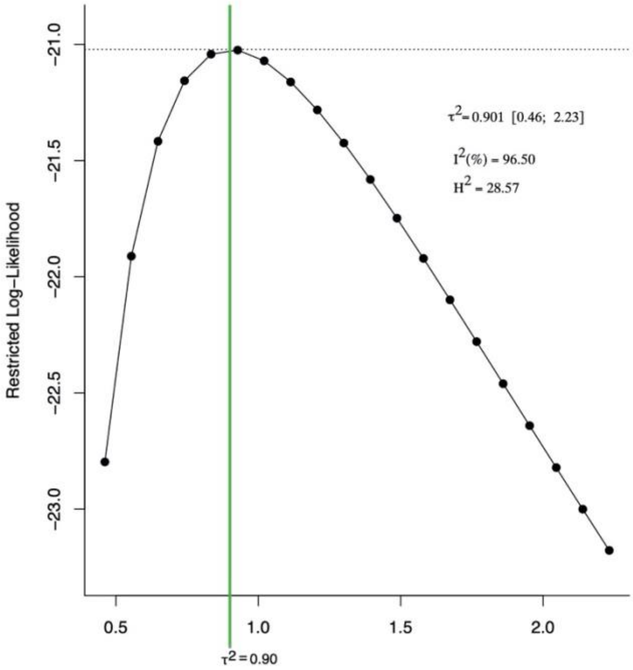


**The dispersion (tau) of the underlying main effect.**

### Supplement 7

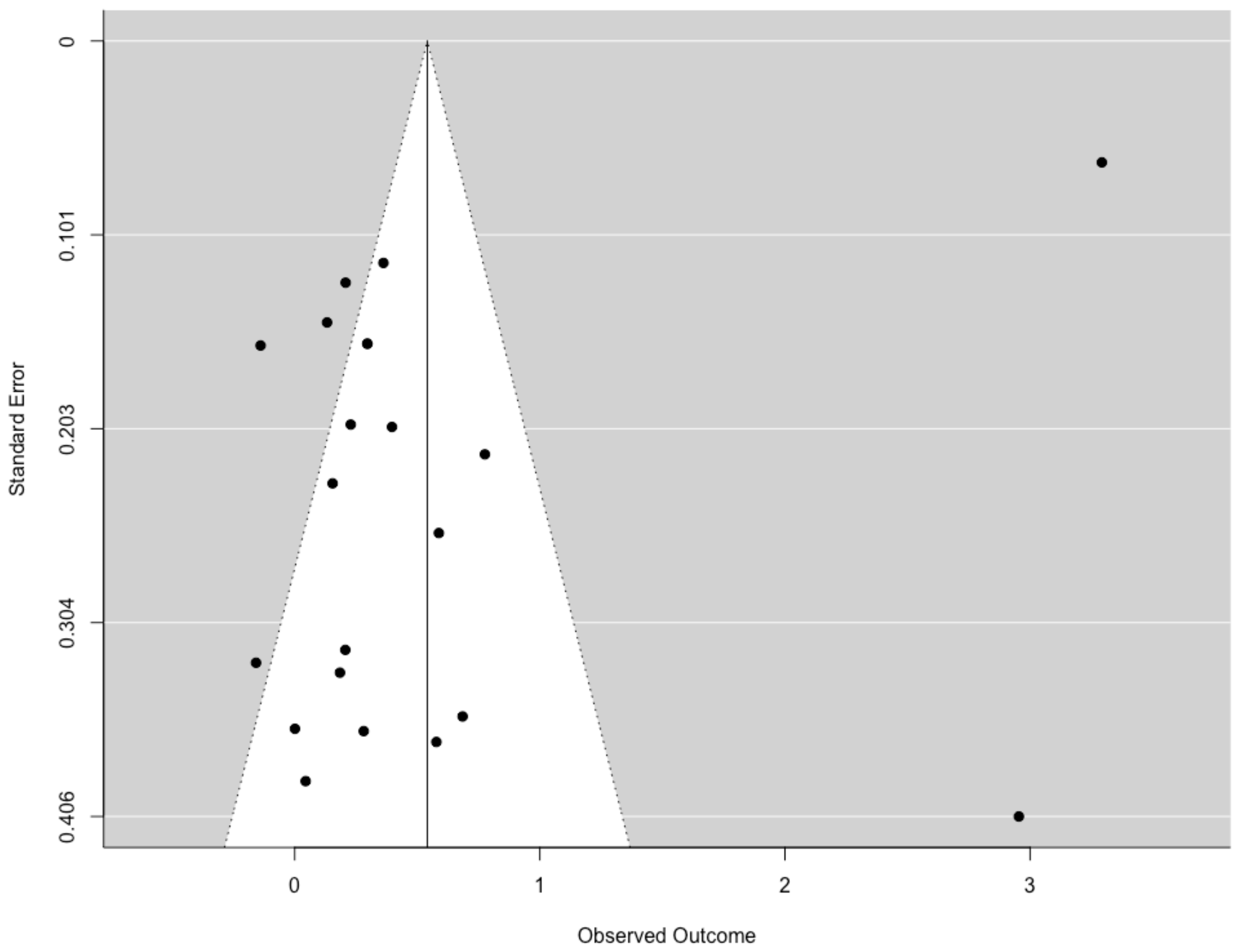
