## Supplement 4 for "Efficacy of interventions to increase physical activity for people with heart failure: a meta-analysis"

**Supplement 4. Expanded Table 2: Intervention characteristics.**

| Author, year | | Intervention description | Behaviour Change Techniques | Intervention intensity | Facilitator | mode of delivery | Theory (TCS) |
|---|---|---|---|---|---|---|---|
| Ajiboye et al., 2015 | *Main intervention* | Aerobic and resistance training and education | BP/R; GT | 36 session; 60-minute sessions; three a week (36 sessions) | nr | face-to-face | none |
| | *Comparator treatment* | usual care and education | PI | | | | |
| Bernocchi et al., 2018 | *Main intervention* | integrated telerehabilitation home-based programme (Telereab- HBP) with personalised exercise maintenance programme | IHC; CS: IHPB; AOE; BP/R; GT; SMB; MbBOwF; FB; | nr | Nurse tutor; physiotherapist tutor | telemonitoring of vital signs. Mini-ergometer, pedometer and diary. | none |
| | *Comparator treatment* | usual care | IHC | | | | |
| van den Berg-Emons et al., 2004 | *Main intervention* | aerobic exercise training | AP; BP/R; GS(B); | 24 sessions, 60-minute sessions twice a week (12 weeks) | not reported | hospital-based training in groups | none |
| | *Comparator treatment* | usual care without particular advice for exercise | none | not reported | not reported | not reported | none |
| Boyne et al., 2014 | *Main intervention* | Individually tailored e-health intervention 'Health Buddy.' | IHC; SMB; SMOB | 364 sessions: daily 10-minute session (52 weeks) | HF nurse and a nurse assistant | Telemonitoring device 'Health Buddy.' | none |
| | *Comparator treatment* | education | IHC | not reported | not reported | Home-based, individual | none |
| Brodie et al., 2005 | *Main intervention* | Motivational Interviewing | GT; IHC; PS; SC; SMB; SS(E); SS(U); | Eight sessions: Weekly 60- minute sessions (8 weeks) | A researcher without clinical qualification | home-based Face-to-face sessions | MI (TCS = 2) |
| | *Main intervention* | Motivational interviewing + education | GT; PS; SC; SMB; SS(E); SS(U) | Eight sessions: Weekly 60- minute sessions (8 weeks) | HF specialist nurse; researcher without clinical qualification | home-based Face-to-face sessions + Usual care package | MI (TCS = 2) |

| Author, year | | Intervention description | Behaviour Change Techniques | Intervention intensity | Facilitator | mode of delivery | Theory (TCS) |
|---|---|---|---|---|---|---|---|
| | Comparator treatment | education | IHC | not reported | HF specialist nurse | Usual care package | none |
| Collins et al., 2004 | *Main intervention* | Aerobic exercise training | AOE; AP; BP/R; GS(B); GT; IHPB; RP/C | 120 sessions five days a week 50 minutes (24 weeks) | Exercise physiologist or nurse | Supervised group-based | none |
| | *Comparator treatment* | usual care | none | not reported | not reported | not reported | none |
| Corvera-Tindel et al., 2004 | *Main intervention* | A home walking exercise programme | BP/R; GS(B); GT; MBbOwF; MOBwF; SMB | 60 sessions: 60 minutes 5 days a week (12 weeks) | nurse | Home-based, Supervised | none |
| | *Comparator treatment* | usual care | MBbOwF | not reported | not reported | not reported | none |
| Cowie et al., 2011 | *Main intervention* | hospital-based aerobic exercise training | Intervention 1: AP; BP/R; DB; GS(B); GT; IHC; IHPB; RBG; SMB; SMOB; | 16 sessions: 60 minutes sessions, Twice a week eight weeks | Exercise instructor | Face-to-face, hospital-based | none |
| | *Main intervention* | home-based exercise training | Intervention 2: AP; BP/R ; DB; GS(B); GT; GTB: IHC; IHPB: SMOB; | 16 sessions: 30 minutes sessions, Twice a week eight weeks | physiotherapist | home-based, individual (DVD) | none |
| | *Comparator treatment* | usual care | none | not reported | not reported | not reported | none |
| Dalal et al., 2018 (REACH-HF) | *Main intervention* | the Rehabilitation Enablement in Chronic Heart Failure (REACH-HF) self-care and rehabilitation intervention | BP/R; RNE; RPE; IHC; SS(E); SS(P); SS(U); GT; GS; PS; RBG; SMB | at least three face-to-face sessions; via phone - unspecified; 12 weeks | Two trained cardiac nurses | nr | SDT, CSM, CT (TCS = 5) |
| | *Comparator treatment* | usual care | IHC | | | | |
| Freedland et al., 2018 | *main intervention* | Integrative Cognitive Behaviour Therapy + Enhanced (with education) usual care | IHC; GS(B); AP; CS; IHPB; PS; MBOwF; MOBwF; ST; PC; | 25 sessions; 60-minute sessions; once a week; 4 education sessions via phone (30 minutes) | Clinical phycology trainee (graduate student) | nr | CBT (TCS = 6) |

| Author, year | | Intervention description | Behaviour Change Techniques | Intervention intensity | Facilitator | mode of delivery | Theory (TCS) |
|---|---|---|---|---|---|---|---|
| | *Comparator treatment* | Enhanced (with education) usual care | IHC | Four education sessions via phone (30 minutes) | Nurse | nr | none |
| O'Connor et al., 2009 (HF-ACTION) | *Main intervention* | Aerobic exercise training + Exercise adherence facilitation intervention | AOE; AP; BP/R; CS; GS(B); GT; GTB; IHC; IHPB; PC; SMB; SMOB; SS(E); MBbOwF; SS(P); SS(U) | 72 sessions, three sessions per week (24 weeks) | Physiotherapist | Facility-based group-based exercise training | TTM, SCT (TCS = 7) |
| | *Comparator treatment* | usual care | CS; GS(B); IHC; MBbOwF; SS(U); | not reported | not reported | not reported | none |
| Jolly et al., 2009 | *Main intervention* | Aerobic and resistance exercise training | AP; BC; BP/R; DB; GS(B); GT; IHC; IHPB SMB; SS(U) | Three supervised exercise sessions; 3 home visits; 3 telephone sessions; 120 self-applied sessions (5 times a week) 20-30 minutes (24 weeks) | PA instructor | Home-based, face-to-face | none |
| | *Comparator treatment* | HF specialist nurse care | IHC | not reported | HF specialist nurse | not reported | none |
| Meng et al., 2016 | *Main intervention* | self-management patient education program + inpatient cardiac rehabilitation | IHC; GS(B); AP; PS; RBG; SMB; FPS; IHPB; BF | Five sessions; 60-75 minutes; 2 sessions a week (approx), three-week session; | Physician; nurse; psychologist; physiotherapist | face-to-face | **nr** |
| | *Comparator treatment* | education | IHC | One session; 60 minutes | physician | face-to-face | |
| Pozehl et al., 2018 (HEART Camp) | *Main intervention* | multicomponent intervention Heart Failure Exercise and Resistance Training (Heart Camp) | IHTB; BP/R; BF; IEC; iHC; MOBwF; SS(U); GS(B); RBG; PS; VPaC; FB; BF; SMB | Six group-based educational sessions (adoption: 6 and months), self-administered (maintenance at 13-18 months) one session a week (18 months) | coach trainer | nr | SCT (TCS = 5) |

| Author, year | | Intervention description | Behaviour Change Techniques | Intervention intensity | Facilitator | mode of delivery | Theory (TCS) |
|---|---|---|---|---|---|---|---|
| | *Comparator treatment* | Enhanced (nine exercise sessions for three months) | IHPB; BP/R | nr | nr | face-to-face | none |
| Smeulders et al., 2009 | *Main intervention* | Chronic disease management programme | AP; BC; BE; BP/R; D; DB; FB; IHC; ISRM; PS; R; RNE; SS(U); ST | Six sessions 150 minutes once a week (6 weeks) | Lay leader (HF patient); HF specialist nurse | Hospital-based group-based exercise training and classes | SLT ( TCS = 8) |
| | *Comparator treatment* | usual care | none | not reported | HF specialist nurse | not reported | none |
| Tomita et al., 2008 | *Main intervention* | Multidisciplinary Internet-based programme on management of HF | FB; IHC; IHPB; SMB | Forty-two sessions, 3.5 sessions a month for about 10 minutes. 1-year e-health intervention | Self-applied (Website) | Home-based, internet-based (website) | TTM, SST (TCS = 2) |
| | *Comparator treatment* | Usual care | none | not reported | not reported | not reported | |
| Willenheimer et al ., 2001 | *Main intervention* | Aerobic exercise training | AP; BP/R; DB; GS(B); GT; IHPB | 41 session: 2 sessions a week (15 minutes) for seven weeks: and then three sessions a week (45 minutes) for nine weeks | physiotherapist | Hospital-based, Group-based exercise training | none |
| | *Comparator treatment* | usual care + discouragement to exercise | PI | 16 weeks | nr | not reported | none |
| Witham et al., 2005 | *Main intervention* | Seated aerobic exercise training followed by seated resistance exercise training | BP/R; GS(B); GT; GTB; IHC; MOBwF; SMB; SS(U) | 17-20 sessions 20-minute session Twice a week (12 weeks) | Physiotherapist | Group-based, hospital-based exercise training (supervised and home settings) Followed by home self-monitoring, self- | none |

| Author, year | | Intervention description | Behaviour Change Techniques | Intervention intensity | Facilitator | mode of delivery | Theory (TCS) |
|---|---|---|---|---|---|---|---|
| | | | | | | monitoring and goal setting. | |
| | *Comparator treatment* | usual care | IHC | nr | nr | not reported | none |
| Yeh et al., 2011 | *Main intervention* | Exercise training (Tai Chi Mind-Body movement) | AP; BC; BP/R; DB; GS(B); IHPB; SMB | twice a week (group sessions); three times a week home sessions) one hour (group sessions); 35 minutes (home sessions) (12 weeks) | Exercise instructor | Hospital-based, Group-based exercise training | none |
| | *Comparator treatment* | usual care | none | Not reported | Video recording | Followed by home-based exercise training and monitoring | none |
| Young et al. 2015; 2016 | *Main intervention* | Patient Activation Intervention on self-management in HF (Patient AcTivated Care at Home: PATCH) | IHC; DB; IHPB; SMOB; ;GS(B); IAwCB; SC; VC; MBbOwF; NSI | 12 sessions (45 minutes); one in a hospital and then twice a week for the first two weeks, once a week for weeks 3–6, and every other week for weeks 7–12 (12 weeks) | Advanced practice nurse | one session face-to-face; telephone | none |
| | *Comparator treatment* | Usual care | IHC | 50-minute one session | nurse | face-to-face | none |

*Note: TCS – Theory Coding Scheme; nr – not reported; MI – Motivational Interviewing, SDT – Self-determination theory; CSM -- CT—Control Theory; CBT—Cognitive Behavioural Therapy; TTM – Transtheoretical Model of Change; SCT—Social Cognitive Theory; SLT – Social Learning theory; SST;* AOE – 12.5. Adding objects to the environment; AP – 1.4. Action planning; BC –12.6. Body changes; BE – 4.4. Behavioural experiments: BP/R – 8.1. Behavioural practice/rehearsal; BF - Biofeedback; CS – 9.2. Credible source; D – 12.4. Distraction; DB – 6.1. Demonstration of the behaviour;FB – 2.2. Feedback on behaviour; FPS - 15.3. Focus on past success; GS(B) – 1.1. Goal setting (behaviour); GT – 8.7. Graded tasks; GTB – 8.6. Generalisation of target behaviour; IEC - 5.6. Information about emotional consequences; IHC – 5.1 Information about health consequences; IHPB – 4.1. Instruction on how to perform the behaviour; IAwCB – 13.5. Identity associated with changed behaviour; ISRM – 13.1. Identification of self as a role model; MBbOwF - 2.5. Monitoring of behaviour by others without feedback; MOBwF – 2.5. Monitoring of outcomes of behaviour without feedback; NSI – non-specific incentive; PC – 7.1. Prompts/cues; PS – 1.2. Problem-solving; R – 4.3. Reattribution; RBG – 1.5. Review behaviour goal(s); RNE –11.2. Reduce negative emotions; RP/C – 7.3. Reduce prompts/cues; RPE - 12.1. Restructuring the physical environment; SC – 6.2. Social comparison; SMB –

2.3. Self-monitoring of behaviour; SMOB – 2.4. Self-monitoring of outcome(s) of behaviour;  SS(E) – 3.3. Social support (emotional); SS(P) –3.2. Social support (practical); SS(U) – 3.1. Social support (unspecified); ST – 15.4. Self-talk;  VPaC - 15.1. Verbal persuasion about capability; VC  - 16.3. Vicarious consequences
