## Supplement 5 for "Efficacy of interventions to increase physical activity for people with heart failure: a meta-analysis"

**Supplement 5.** Non-significant results: Intervention and participant characteristics, as well as method of assessment that do not contribute to intervention efficacy.

| Variable | SMD/b | CI, 95% (lower) | CI, 95% (upper) |
|---|---|---|---|
| ***Theory use:*** | | | |
| Theory use score (overall, TCS) | 0.13 | -0.01 | 0.27 |
| Theory mentioned | 0.46 | -0.23 | 1.15 |
| ***Behaviour Change Techniques:**** | | | |
| Self-monitoring of behavior | 0.49 | -0.04 | 1.03 |
| Information about health consequences | 0.50 | -0.05 | 1.06 |
| Social support (emotional) | 0.92 | -0.08 | 1.92 |
| Instruction on how to perform a behavior | 0.51 | -0.11 | 1.13 |
| Social support (unspecified) | 0.53 | -0.19 | 1.25 |
| Demonstration of the behavior | 0.37 | -0.39 | 1.12 |
| Feedback on behavior | 0.31 | -0.77 | 1.38 |
| Problem solving | 0.05 | -0.92 | 1.02 |
| Monitoring outcome(s) of behavior by others without feedback | 0.32 | -0.92 | 1.56 |
| Behavioral contract | 0.30 | -0.93 | 1.53 |
| Biofeedback | 0.50 | -1.01 | 2.01 |
| Review behavior goal(s) | 0.18 | -1.08 | 1.43 |
| Reduce negative emotions | 0.07 | -1.17 | 1.30 |
| Information about emotional consequences | 0.04 | -1.47 | 1.56 |
| Vicarious consequences | 0.23 | -1.47 | 2.39 |
| Self-talk | 0.04 | -1.48 | 1.55 |
| Social comparison | 0.02 | -1.55 | 1.60 |
| Behavioral experiments | 0.21 | -1.93 | 2.34 |
| Distraction | 0.21 | -1.93 | 2.34 |
| Identification of self as role model | 0.21 | -1.93 | 2.34 |
| Reattribution | 0.21 | -1.93 | 2.34 |
| Identity associated with changed behaviour | 0.23 | -1.93 | 2.39 |
| Non-specific incentive | 0.23 | -1.93 | 2.39 |
| Restructuring the physical environment | 0.13 | -2.01 | 2.27 |
| Reduce prompts/cues | 0.04 | -2.22 | 2.31 |
| Social support (practical) | 1.42 | -2.22 | 2.46 |
| Discrepancy between current behavior and goal | -0.14 | -2.28 | 2.01 |
| ***Intervention intensity:*** | | | |
| Number of sessions | 0.00 | -0.01 | 0.01 |

| | | | |
|---|---|---|---|
| Duration, single session (mins) | 0.00 | 0.00 | 0.01 |
| Intervention contact time (mins) | 0.00 | 0.00 | 0.00 |
| intervention duration (weeks) | -0.01 | -0.04 | 0.02 |
| Individually delivered | 0.30 | -0.47 | 1.06 |
| ***Setting:*** | | | |
| Home-setting | 0.16 | -0.66 | 0.99 |
| ***Facilitator:*** | | | |
| Telehealth | 0.73 | -0.79 | 2.25 |
| Nurse | 0.34 | -0.38 | 1.05 |
| HF nurse | 0.09 | -1.18 | 1.36 |
| Researcher | 0.29 | -1.25 | 1.82 |
| Self-applied | 0.21 | -1.35 | 1.76 |
| Lay leader | 0.21 | -1.93 | 2.34 |
| Advanced practice nurse | 0.23 | -1.93 | 2.39 |
| Cardiac nurse | 0.13 | -2.01 | 2.27 |
| Website | 0.00 | -2.25 | 2.25 |
| Graduate student therapist (trained) | -0.14 | -2.28 | 2.01 |
| ***Participant characteristics:*** | | | |
| Age | 0.01 | 0.00 | 0.01 |
| Males included in the sample, (%) | 0.01 | 0.00 | 0.02 |
| LVEF, (%) | 0.01 | 0.00 | 0.02 |
| ***Physical activity assessment:*** | | | |
| Self-reports | 0.68 | -0.06 | 1.41 |
| Accelerometer | 0.31 | 0.07 | 0.56 |
| Difference in efficacy between trials using accelerometers vs self-reports | -0.32 | -1.15 | 0.51 |

*(BCTTV1 taxonomy: Michie et al., 2013)
