## Supplement 6 for "Efficacy of interventions to increase physical activity for people with heart failure: a meta-analysis"

**Supplement 6: The sensitivity analysis results.**

| EXCLUDED TRIALS | Overall effect | General Approach | Efficacious intervention characteristics |
|---|---|---|---|
| Trials with the education comparator (Boyne et al. 2014; Meng et al. 2016; Freedland et al. 2015; Brodie and Inoue 2005) (Ajiboye et al. 2015) | SMD = 0.31, 95%CI [0.21; 0.40] *** | **Exercise: SMD = 0.34 , 95%CI [0.18; 0.51];** **Remote communication and treatment: SMD = 0.42, 95%CI [0.24; 0.60];** Exercise and behaviour change: SMD = 0.21, 95%CI [ 0.004; 0.41]; Disease Management: SMD = 0.21, 95%CI [ -0.03; 0.45]; Self-Management: SMD = 0.23, 95%CI [-0.17; 0.62]. | No change in the significance of the effects associated with individual intervention or participant characteristics |

| EXCLUDED TRIALS | Overall effect | General Approach | Efficacious intervention characteristics |
|---|---|---|---|
| HF-ACTION trial<br>(O’Connor et al. 2009) | SMD = 0.37,<br>95% CI: [0.10; 0.63]* | **Exercise: SMD = 0.56, 95%CI [0.18; 0.94];**<br>Remote communication and treatment:<br>SMD =0.41, 95%CI [-0.29; 1.11];<br>MI: SMD = 0.03, 95%CI [-0.89; 0.94];<br>**Exercise and behaviour change:**<br>**SMD = 0.21, 95%CI [ -0.59; 1.01];**<br>CBT: SMD = 0.21, 95%CI [-0.91; 1.33];<br>Disease Management: SMD =0.23, 95%CI [-0.95; 1.41]. | Behavioural Practice and Rehearsal *<br>Graded task *<br>Group-based *<br>Centre-based * |

| | | | |
|---|---|---|---|
| High Risk of Bias trials<br>(Ajiboye et al. 2015;<br>Tomita et al. 2008) | SMD =<br>0.4578 95%CI<br>[0.0903<br>0.8252 * | Exercise: SMD =0.36, 95%CI [-0.2; 0.91];<br>Remote communication and treatment:<br>SMD = 0.56, 95%CI [-0.57; 1.7];<br>MI: SMD = 0.03, 95%CI [ -1.16; 1.21];<br>Exercise and behaviour change:<br>SMD = 1.27, 95%CI [0.47; 2.07]***;<br>CBT: SMD = 0.21, 95%CI [ -1.38; 1.8];<br>Disease Management: SMD =0.23, 95%CI [ -1.39 1.85]. | **Social Support (emotional)** *<br>**Social Support (practical)** **<br>**TCS score** *<br>Monitoring of behaviour by others w/o feedback*<br>Credible source***<br>Adding objects to the environment ***<br>Self-monitoring of outcome(s) of behaviour*<br>**Self-monitoring of behaviour** *<br>**Information about health consequences** *<br>**Information on how to perform behaviour** *<br>Graded tasks *<br>Action Planning *<br>Goal setting (behaviour) *<br>Behavioural Practice and Rehearsal** |

| EXCLUDED TRIALS | Overall effect | General Approach | Efficacious intervention characteristics |
|---|---|---|---|
| HF-ACTION trial<br>(O'Connor et al. 2009)<br>and High Risk of Bias<br>trials | SMD = 0.2670<br>95%CI [0.1549<br>0.3790 *** | **Exercise: SMD=0.35, 95% CI [0.15; 0.54]*;**<br>**Remote communication and treatment:**<br>**SMD=0.47, 95%CI [0.24;0.70]*;**<br>MI: SMD=0.03, 95%CI [ -0.46;0.52];<br>**Exercise and behaviour change:**<br>**SMD= 0.21; 95%CI:[-0.07; 0.49];**<br>CBT SMD= 0.21, 95%CI:[-0.15; 0.57]<br>Disease Management SMD= 0.23, 95%CI:[-0.25; 0.71]. | **Nurse** ***<br>**Telehealth****<br>**Biological Feedback***<br>Credible source *<br>Self-monitoring of outcomes of behaviour*<br>**Self-monitoring of behaviour** **<br>**Demonstration of the behaviour** **<br>**Information about health consequences** **<br>Graded tasks **<br>Action Planning **<br>Goal setting (behaviour)***<br>Behavioural Practice and Rehearsal*** |

**Note: *p<0.05; **p<0.01,*** p<0.001. The changes in the findings, identified by the sensitivity analysis (corresponding exclusion of the trials) are in bold.**
